# Exploring the power of MRI and clinical measures in predicting Alzheimer’s disease neuropathology

**DOI:** 10.1101/2024.07.09.24310163

**Authors:** Farooq Kamal, Cassandra Morrison, Michael D. Oliver, Mahsa Dadar

## Abstract

**Background:** The ability to predict Alzheimer’s disease (AD) before diagnosis is a topic of intense research. Early diagnosis would aid in improving treatment and intervention options, however, there are no current methods that can accurately predict AD years in advance. This study examines a novel machine learning approach that integrates the combined effects of vascular (white matter hyperintensities, WMHs), and structural brain changes (gray matter, GM) with clinical factors (cognitive status) to predict post-mortem neuropathological outcomes.

**Methods:** Healthy older adults, participants with mild cognitive impairment, and AD from the Alzheimer’s Disease Neuroimaging Initiative dataset with both post-mortem neuropathology data and antemortem MRI and clinical data were included. Longitudinal data were analyzed across three intervals before death (post-mortem data): 0-4 years, 4-8 years, and 8-14 years. Additionally, cross-sectional data at the last visit or interval (within four years, 0-4 years) before death were also examined. Machine learning models including gradient boosting, bagging, support vector regression, and linear regression were implemented. These models were applied towards feature selection of the top seven MRI, clinical, and demographic data to identify the best performing set of variables that could predict postmortem neuropathology outcomes (i.e., neurofibrillary tangles, neuritic plaques, diffuse plaques, senile/amyloid plaques, and amyloid angiopathy).

**Results:** A total of 94 participants (55-90 years of age) were included in the study. At last visit, the best-performing model included total and temporal lobe WMHs and achieved *r*=0.87(*RMSE*=0.62) during cross-validation for neuritic plaques. For longitudinal assessments across different intervals, the best-performing model included regional GM (i.e., hippocampus, amygdala, caudate) and frontal lobe WMH and achieved *r*=0.93(*RMSE*=0.59) during cross-validation for neurofibrillary tangles. For MRI and clinical predictors and clinical-only predictors, *t*-tests demonstrated significant differences at all intervals before death (*t*[-13.60-7.90], *p*-values<0.001). Overall, post-mortem neuropathology outcome were predicted up to 14 years before death with high accuracies (∼90%).

**Conclusions:** Prediction accuracy was higher for post-mortem neuropathology outcomes that included MRI (WMHs, GM) and clinical features compared to clinical-only features. These findings highlight that MRI features are critical to successfully predict AD-related pathology years in advance which will improve participant selection for clinical trials, treatments, and intervention options.

## Background

Alzheimer’s disease (AD) is a neurodegenerative disease characterized by a progressive decline in cognitive functioning. AD is characterized by pathological accumulation of neurofibrillary tau tangles and amyloid-beta deposits [1]. In addition to these pathological changes, structural brain alterations, including gray matter (GM) and white matter (WM) atrophy, and white matter hyperintensity (WMH) burden contribute to cognitive decline in AD [2–5]. For example, GM changes (particularly volume reduction) are closely associated with cognitive impairment in AD, highlighting their importance in the neurodegeneration process [6–8]. Research has also shown a strong relationship between WM volume changes and cognitive impairment across the spectrum of AD progression [9]. Similarly, WMHs are markers of cerebrovascular disease burden and are increasingly recognized for their role in cognitive decline and the progression of neurodegenerative diseases [10–12].

People with AD often present with both structural and cerebrovascular changes such as GM and WM volume loss and WMH burden at autopsy [13–16]. These volumetric changes are commonly studied in the entorhinal cortex [6] and hippocampus [7,8] as they are two of the initial brain regions to show abnormal volume loss in AD, thus providing information about AD staging/progression. Although neurodegeneration has been included as a marker for staging AD, the Alzheimer’s Association Workgroup has recently proposed a revised criteria for diagnosing and staging of AD. The updated guidelines now also include vascular brain injury (often measured using MRI markers of cerebrovascular disease) as a common non-AD co-pathology [1], further highlighting the prevalence of vascular pathology in AD. Taking markers of both structural and cerebrovascular pathology into consideration, the present study employs a novel approach to examine whether these in vivo abnormalities are predictive of neuropathology severity.

Previous neuropathological studies have provided insights into early risk factors for AD by exploring the correlations between neuropathological and clinical or brain volume measures individually. For instance, Kaur et al. [17] explored the association between neuropathology (e.g., neurofibrillary tangle burden (NFT)) and total and regional brain volume and reported significant correlations between neuropathological measures and brain volume. Additionally, a recent study by Zhang et al. [18] further explored the relationship between clinical and neuropsychological measures and neuropathological data in cognitively healthy older adults. Neuropathological measures such as Braak-NFT stage were significantly correlated with clinical measures of memory, executive function, and visuospatial function. These studies predominantly concentrate on either clinical or MRI-based markers that correlate with neuropathology [17–20]. However, these studies did not investigate the potential of using a combination of WMH, WM, and GM volumes as features to predict neuropathology. Thus, a comprehensive approach that combines these features to predict neuropathological outcomes remains unexplored.

The integration of machine learning (ML) has opened new avenues for understanding and diagnosing neurodegenerative diseases. With their ability to handle large datasets and complex nonlinear patterns, ML models have been widely utilized for disease diagnosis and prognosis applications [21]. For example, support vector machines (SVMs) have been used to classify different stages of AD using MRI data, helping to outline disease progression [22]. Similarly, linear regression has been instrumental in identifying biomarkers and predicting disease outcomes in neurodegenerative conditions [18]. Despite these advances, accurately predicting neuropathology remains a significant challenge that has not been previously addressed. Early prediction of neuropathology remains an essential component in AD research because it would aid in improving selection of participants for clinical trials and enhance treatment options. For example, AD-related brain changes occur 10-20 years before diagnosis occurs, and currently there are no methods that can accurately predict AD that early. If researchers develop a method that could predict who would progress to AD, it may be possible to intervene earlier in the disease course and prevent the irreversible brain changes that occur.

Current research has yet to fully incorporate vascular (WMHs) and structural brain changes (WM and GM), with clinical factors in ML models for neuropathology prediction. Given the new criteria for diagnosis and staging^1^, this gap underscores a critical area for exploration, as WMHs represent a potentially valuable marker for early disease detection that is yet to be fully examined. Given the limitations of the existing research, the present study aims to integrate these features with other AD-related brain measures to predict post-mortem neuropathology years in advance. Additionally, the study will compare the importance of MRI measures in addition to clinical measures. To achieve this, ML methods will be applied to determine the importance of different features (e.g., cognitive, regional and global brain volumes, and WMHs) in predicting AD-related neuropathologies. We hypothesize that in addition to typical MRI measures (i.e., brain volume) such as WM and GM, WMH burden will also be a critical feature that contributes to AD-related post-mortem neuropathology prediction.

## Methods

### Participants

Data were obtained from the Alzheimer’s Disease Neuroimaging Initiative (ADNI) database (adni.loni.usc.edu). Participants were included if they had post-mortem neuropathology information, MRIs from which regional white and gray matter and WMH volume could be extracted, completed the Clinical Dementia Rating-Sum of Boxes (CDR-SB, a measure of global cognition), digit span (as a measure of working memory), trail making test - part B (as a measured of processing speed), everyday cognition scale (as a measure of subjective cognitive decline), and logical memory (as a measure of episodic memory). MRI, clinical, and demographic data were collected from participants over time (i.e., longitudinally) before their death (when post-mortem neuropathology information was obtained). A total of 94 participants (55-90 years of age) were included in the study. There were 59 participants with data within 4 years prior to death, 63 participants with data from 4 to 8 years before death, and 28 participants with data between 8 to 14 years before death. Summary of demographic information for participants in ADNI are presented in Table 1.

**Table 1:** Demographic Information in the ADNI dataset across time of death intervals prior to post-mortem.

|  | Time |  |  |
| --- | --- | --- | --- |
|  | *0-4 Years* | *4-8 Years* | *8-14 Years* |
| Sample Size | 59 | 63 | 28 |
| Age | 81.2 ± 6.7 | 82.8 ± 6.9 | 85.4 ± 6.7 |
| Sex (Female) | 15 (25%) | 16 (25%) | 7 (25%) |
| Education | 16.3 ± 2.8 | 16.3 ± 2.8 | 16.6 ± 3.3 |
| Diagnosis |  |  |  |
| NC | 7 (11%) | 7 (11%) | 6 (21%) |
| MCI | 8 (14%) | 26 (41%) | 11 (39%) |
| AD | 44 (75%) | 29 (46%) | 11 (39%) |

Numbers are presented as total (percentage of group) or mean ± standard deviation. NC=cognitively healthy older adults. MCI=mild cognitive impairment. AD = Alzheimer's Disease.

### Structural MRI acquisition and processing of WMH in ADNI

Scans were downloaded from the ADNI website (see http://adni.loni.usc.edu/methods/mri-tool/mri-analysis/ for MRI acquisition protocol). T1w scans were pre-processed through our standard pipeline including noise reduction [23], intensity inhomogeneity correction [24], and intensity normalization into range [0–100]. The pre-processed images were linearly (9 parameters: 3 translation, 3 rotation, and 3 scaling) [25] registered to the MNI-ICBM152-2009c average [26].

A previously validated WMH segmentation technique that has been extensively tested in aging and neurodegenerative diseases was used to obtain WMH measurements [27]. This technique has been previously employed in other multi-center studies[28,29] and has also been validated in the ADNI cohort [30]. The automated WMH segmentation technique extracts a set of location (i.e. spatial priors) and intensity (distribution histograms) features and uses them in combination with a random forest classifier to detect the WMHs in new images [27, 28, 29]. The method has been trained based on a library of manually segmented scans from the ADNI data (50 participants independent of the ones studied here), ensuring its accuracy when applied to the current dataset. Automatic segmentation of the WMHs was completed using only the T1w contrasts. The decision to use only T1w images for WMH segmentation was made since ADNI1 study only includes T1w, and T2w/PD images, whereas ADNI2/GO includes T1w and axial 2D FLAIR images, and ADNI3 includes T1w sagittal 3D FLAIRs. Since the resolution of the images (i.e., voxel sizes) were very different between T2w/PD (1*1*3 mm^3^) and FLAIRs (0.85*0.85m*5 mm^3^ and 1*1*1.2 mm^3^), to be able to reliably combine and use WMH volumes from all these studies, we only used the T1w images for WMH segmentation. We have previously validated the performance of our pipeline in detecting WMHs based on T1w images both in ADNI and another independent dataset, and have shown that the T1w-based WMH volumes (1) hold strong correlations with T2w/PD and FLAIR based WMH volumes (*r*LJ=LJ.89, *p*LJ<LJ.001 for ADNI1, and *r*LJ=LJ.97, *p*LJ<LJ.001 for ADNI2/GO), and (2) have similar relationships with clinical/cognitive scores and risk factors associated with WMHs. ^34^ Finally, the quality of all WMH segmentations was visually assessed by M.D. (blinded to clinical diagnosis). WMH load was defined as the volume of all voxels identified as WMH in the standard space (in mm^3^) and were normalized for head size. Regional (frontal, temporal, occipital, and parietal) and total WMH volumes were calculated based on Hammers Atlas [27,31]. All WMH volumes were also log-transformed to achieve normal distribution.

### Freesurfer Measurements

T1w images were processed using FreeSurfer and quality controlled by the UCSF group, and total WM as well as total and regional GM volumes were extracted. 1.5T and 3T data were processed with FreeSurfer versions 4.3 and 5.1, respectively, as appropriate [32]. Regional GM features examined included the hippocampus, entorhinal cortex, parahippocampal gyrus, fusiform gyrus, and middle temporal gyrus, amygdala, putamen, caudate, and thalamus volumes.

### Predictive modelling with multiple feature sets in ADNI

Analyses were performed using Python software version 3.11. The predictor variables included global cognition, episodic memory, subjective cognitive decline (SCD), perceptual speed, perceptual orientation, working memory, and structural brain volumes (WM, total and regional GM and WMHs). These predictors were obtained based on the last available visit, within 4 years prior to death (0-4 years) and therefore included 1 timepoint per person. Neurofibrillary tangles (classified by Braak stage), neuritic plaques, and diffuse plaques (classified according to CERAD), amyloid plaques (classified according to Thal amyloid phase), and amyloid angiopathy were the outcomes of interest for the prediction models.

All continuous values were standardized using the *StandardScaler* from the *sklearn.preprocessing* package in Python prior to analyses, aligning with best practices in machine learning research [33–35]. We incorporated a range of linear and nonlinear machine learning models, including gradient boosting classifier and regressor, support vector machines, linear regression, and bagging techniques. These models were selected due to their robustness in handling diverse datasets. To evaluate the model performance, we implemented a 10-fold cross-validation (CV) scheme using KFold from *sklearn.model_selection*, minimizing bias and overfitting [36]. Each model within our framework was trained on a subset of the predictors, ranging from individual features to combinations of up to seven features. We evaluated models based on their performance in predicting continuous outcomes. For continuous outcomes, we measured root mean square error (RMSE) and correlation. These metrics provided a comprehensive assessment of model performance.

We examined combinations of predictors using *itertools.combinations*, testing each possible combination to identify the most informative feature set for predicting neuropathology outcomes. To further confirm the validity of our initial results, we employed a bootstrap method, which involved resampling our dataset and performing the same k-fold evaluation on each bootstrap sample. We conducted 50 bootstrap iterations, each involving a 10-fold cross-validation process. This technique, implemented using the resample function, is increasingly recognized for its efficacy in assessing model uncertainty and variance [37,38].

Regression plots were used to visualize and evaluate the predictive performance of machine learning models by comparing neuropathology outcome values against the cross-validated predicted neuropathology values. Higher R^2^ values indicated a better fit of the model to the data, while lower root mean squared error (RMSE) values suggested higher accuracy in predictions.

Longitudinal data at different time intervals before death (0-4 years, 4-8 years, and 8-14 years) were also examined for all predictor variables across the different neuropathological outcomes to investigate the capabilities of the ML models in predicting future pathology. Longitudinal data included multiple timepoints per person. To evaluate model performance in the longitudinal data, we utilized grouped k-fold cross-validation, which stratifies the data based on participants. This method ensures that the same participant does not appear in both training and test sets within a fold, thereby preventing data leakage and ensuring robust model evaluation. Similar modelling procedures were used to examine the longitudinal data. T-tests were used to assess whether the model performances were statistically different across time-to-death groups. The results were corrected for multiple comparisons using false discovery rate (FDR) controlling method.

## Results

The most informative predictors of neuropathology outcomes across each model are presented in Table 2.

**Table 2:**
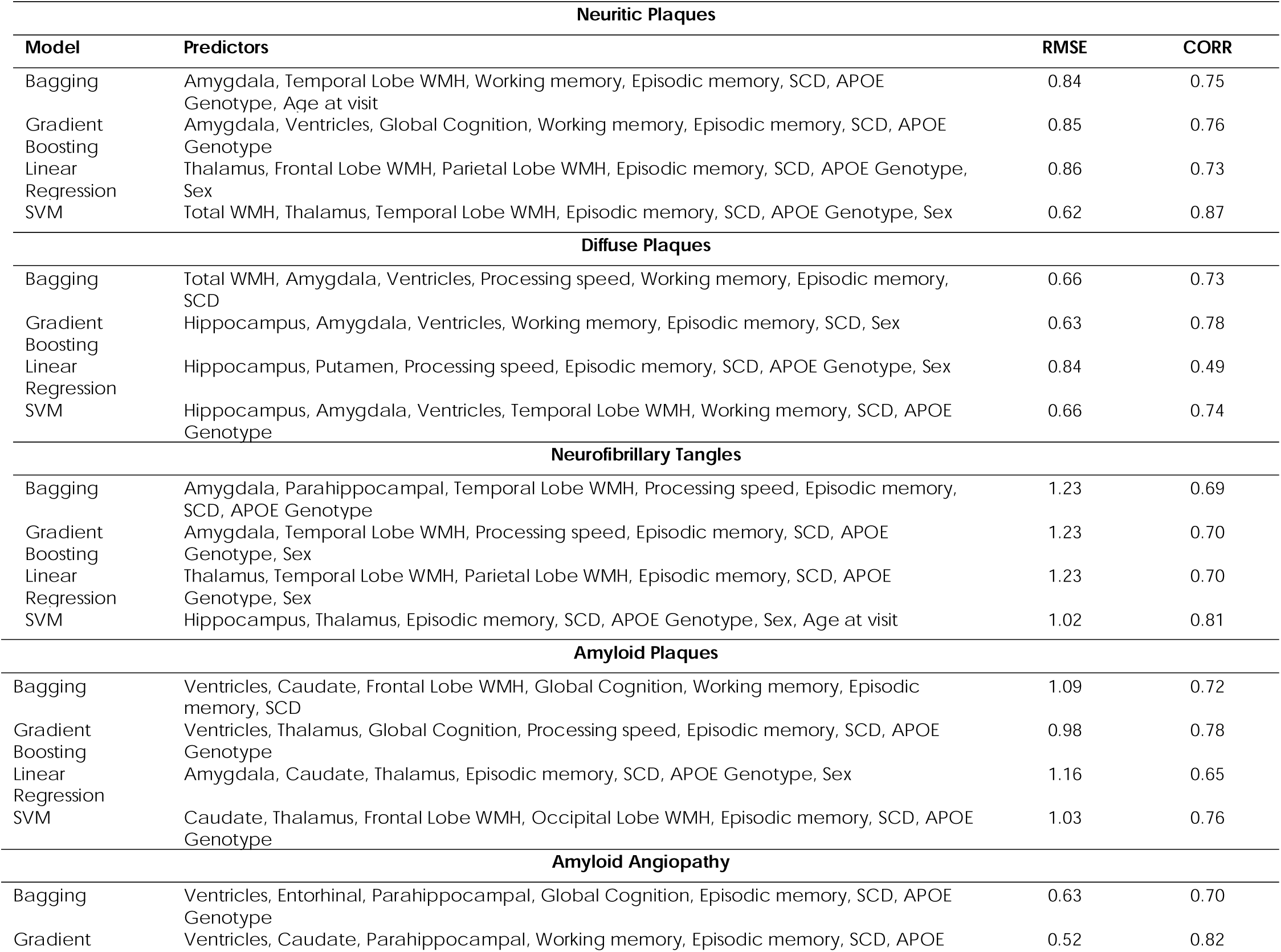
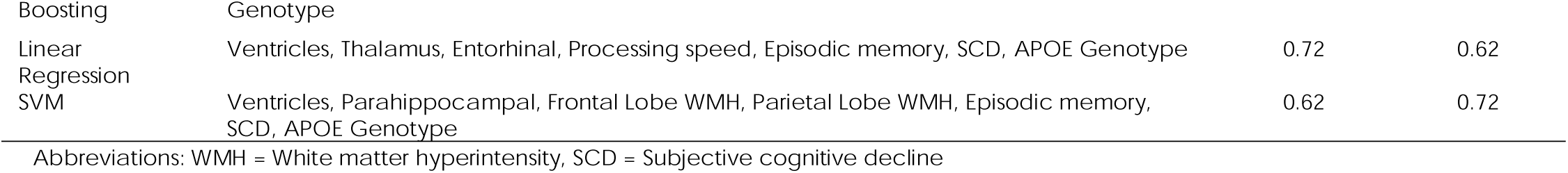
MRI and clinical predictors of neuropathology outcomes across each model at last visit (within 4 years prior to death)

### Neuropathology Prediction

For neuritic plaques, the SVM model, which included total WMH, Thalamus, Temporal Lobe WMH, Episodic memory, SCD, APOE genotype, and Sex, achieved the highest correlation of 0.87 with an RMSE of 0.62 (Fig. 1A). This model outperformed others in predictive accuracy for neuritic plaques. The Gradient Boosting model included predictors such as Amygdala, Ventricles, Global Cognition, Working memory, Episodic memory, SCD, and APOE genotype, demonstrating a high correlation of 0.76 with the lowest RMSE of 0.85. The Bagging model, incorporating predictors like Amygdala, Temporal Lobe WMH, Working memory, Episodic memory, SCD, APOE genotype, and Age at visit, also showed good predictive performance with a correlation of 0.75 and an RMSE of 0.84. In contrast, the Linear Regression model, which included the Thalamus, Frontal Lobe WMH, Parietal Lobe WMH, Episodic memory, SCD, APOE Genotype, and Sex, had a lower correlation of 0.73 and a higher RMSE of 0.86.

**Figure 1:**
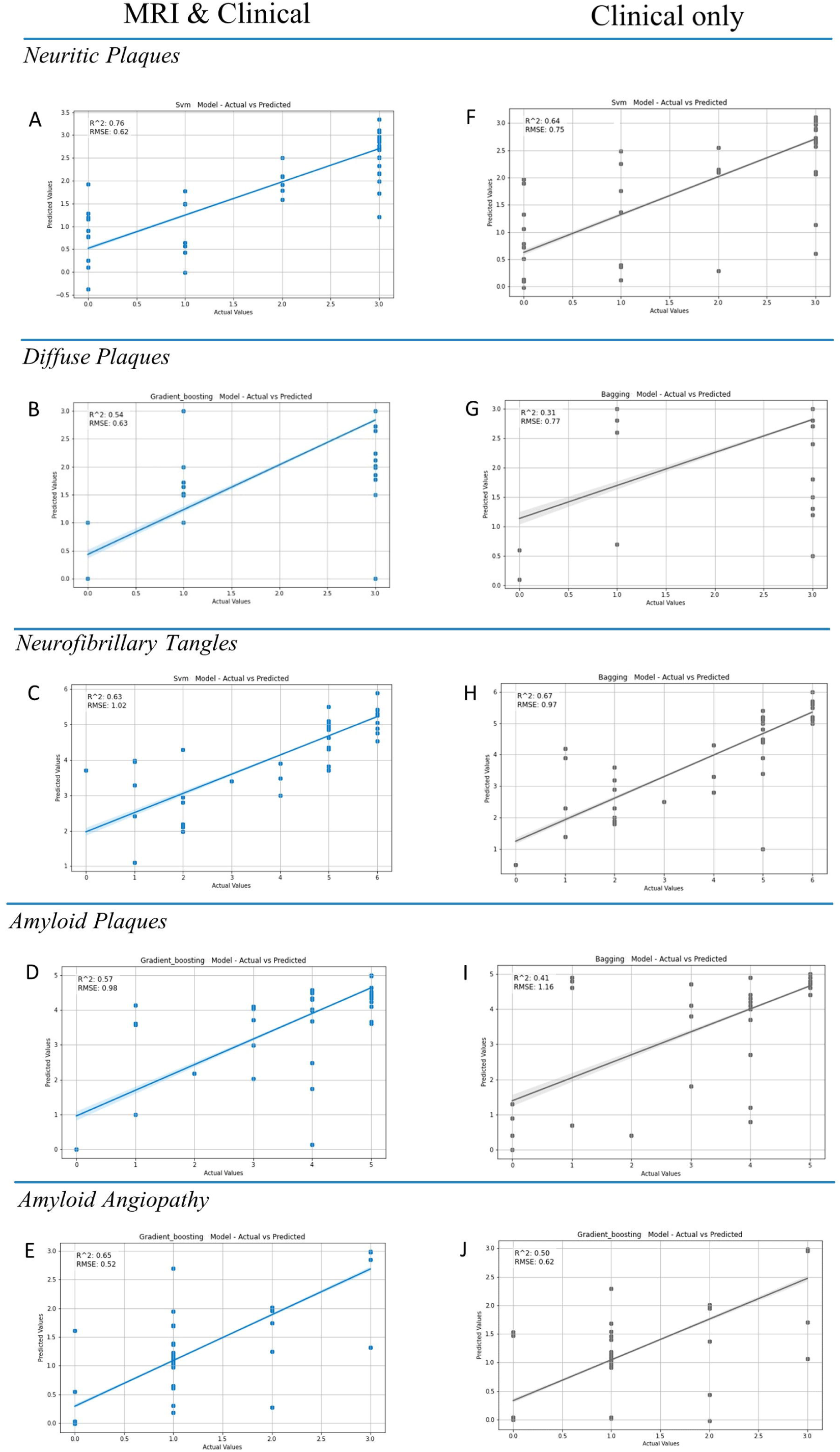
Scatter plots showing the performance of the best models across post-mortem neuropathologies. The models compare actual versus predicted values for neuropathological outcomes across MRI and Clinical (**A-E**), and Clinical only (**F-J**). The regression line represents the best linear fit through the data points. The shaded area around the regression line represents the 95% confidence interval.

In predicting diffuse plaques, the Gradient Boosting model used Hippocampus, Amygdala, Ventricles, Working memory, Episodic memory, SCD, and Sex, showing the strongest performance with an RMSE of 0.63 and a correlation of 0.78 (Fig. 1B). The Bagging model included Total WMH, Amygdala, Ventricles, Processing speed, Working memory, Episodic memory, and SCD, achieving an RMSE of 0.66 and a correlation of 0.73. The Linear Regression model incorporated the Hippocampus, Putamen, Processing speed, Episodic memory, SCD, APOE Genotype, and Sex, with an RMSE of 0.84 and a correlation of 0.49. The SVM model, with Hippocampus, Amygdala, Ventricles, Temporal Lobe WMH, Working memory, SCD, and APOE genotype, showed an RMSE of 0.66 and a correlation of 0.74.

For neurofibrillary tangles, the Gradient Boosting model included Amygdala, Temporal Lobe WMH, Processing speed, Episodic memory, SCD, APOE genotype, and Sex, achieving a correlation of 0.70 and an RMSE of 1.23. The Bagging model included the Amygdala, Parahippocampal, Temporal Lobe WMH, Processing speed, Episodic memory, SCD, and APOE genotype, with a correlation of 0.69 and an RMSE of 1.23. The Linear Regression model incorporated the Thalamus, Temporal Lobe WMH, Parietal Lobe WMH, Episodic memory, SCD, APOE genotype, and Sex, showing a correlation of 0.70 and the same RMSE of 1.23. The SVM model incorporated Hippocampus, Thalamus, Episodic memory, SCD, APOE genotype, Sex, and Age at visit, achieving a correlation of 0.81 and an RMSE of 1.02 (Fig. 1C).

In predicting amyloid plaques, the Gradient Boosting model included the Ventricles, Thalamus, Global Cognition, Processing speed, Episodic memory, SCD, and APOE genotype, achieving a correlation of 0.78 and an RMSE of 0.98 (Fig. 1D). The Bagging model incorporated the Ventricles, Caudate, Frontal Lobe WMH, Global Cognition, Working memory, Episodic memory, SCD, achieving a correlation of 0.72 and an RMSE of 1.09. The Linear Regression model included Amygdala, Caudate, Thalamus, Episodic memory, SCD, APOE genotype, and Sex, resulting in a correlation of 0.65 and an RMSE of 1.16. The SVM model incorporated Caudate, Thalamus, Frontal Lobe WMH, Occipital Lobe WMH, Episodic memory, SCD, and APOE genotype, with a correlation of 0.76 and an RMSE of 1.03.

For amyloid angiopathy, the Gradient Boosting model included the Ventricles, Caudate, Parahippocampal, Working memory, Episodic memory, SCD, and APOE genotype, achieving the best correlation of 0.82 and the lowest RMSE of 0.52 (Fig. 1E). The Bagging model included Ventricles, Entorhinal, Parahippocampal, Global cognition, Episodic memory, SCD, and APOE genotype, with a correlation of 0.70 and an RMSE of 0.63. The Linear Regression model incorporated the Ventricles, Thalamus, Entorhinal, Processing speed, Episodic memory, SCD, and APOE genotype, showing a correlation of 0.62 and an RMSE of 0.72. The SVM model employed Ventricles, Parahippocampal, Frontal Lobe WMH, Parietal Lobe WMH, Episodic memory, SCD, and APOE genotype, with a correlation of 0.72 and an RMSE of 0.62.

### Neuropathology Prediction with clinical predictors

To examine whether MRI features enhance predictive performance beyond that achieved with clinical features alone, we conducted a subsequent analysis focusing solely on clinical features (see Table 3). For neuritic plaques, the SVM model, with clinical predictors such as Global Cognition, Working memory, Episodic memory, SCD, APOE genotype, Sex, and Age at visit, demonstrated the highest correlation of 0.80 and the lowest RMSE of 0.75 (Fig 1F). The Bagging model, with a similar set of predictors including Global Cognition, Processing speed, Working memory, SCD, APOE genotype, Sex, and Age at visit, also showed a high correlation of 0.79 but with a slightly higher RMSE of 0.78. The Gradient Boosting model, incorporating Processing speed, Working memory, Episodic memory, APOE genotype, Sex, and Age at visit, achieved a correlation of 0.74 and an RMSE of 0.89. The Linear Regression model, which included Global cognition among other cognitive and genetic factors, showed a lower correlation of 0.72 and a higher RMSE of 0.88.

**Table 3:** Clinical predictors of neuropathology outcomes across each model at last visit (within 4 years prior to death)

| Neuritic Plaques |  |  |  |
| --- | --- | --- | --- |
| Model | Predictors | RMSE | CORR |
| Bagging | Global Cognition, Processing speed, Working memory, SCD, APOE Genotype, Sex, Age at visit | 0.78 | 0.79 |
| Gradient Boosting | Global Cognition, Processing speed, Working memory, Episodic memory, APOE Genotype, Sex, Age at visit | 0.89 | 0.74 |
| Linear Regression | Global Cognition, Working memory, Episodic memory, SCD, APOE Genotype, Sex, Age at visit | 0.88 | 0.72 |
| SVM | Global Cognition, Working memory, Episodic memory, SCD, APOE Genotype, Sex, Age at visit | 0.75 | 0.80 |
| Diffuse Plaques |  |  |  |
| Bagging | Global Cognition, Processing speed, Working memory, Episodic memory, SCD, APOE Genotype, Sex | 0.77 | 0.62 |
| Gradient Boosting | Global Cognition, Processing speed, Working memory, Episodic memory, SCD, Sex, Age at visit | 0.85 | 0.59 |
| Linear Regression | Global Cognition, Processing speed, Working memory, Episodic memory, SCD, APOE Genotype, Age at visit | 0.89 | 0.44 |
| SVM | Global Cognition, Processing speed, Working memory, SCD, APOE Genotype, Sex, Age at visit | 0.78 | 0.55 |
| Neurofibrillary Tangles |  |  |  |
| Bagging | Processing speed, Working memory, Episodic memory, SCD, APOE Genotype, Sex, Age at visit | 0.97 | 0.82 |
| Gradient Boosting | Processing speed, Working memory, Episodic memory, SCD, APOE Genotype, Sex, Age at visit | 1.14 | 0.74 |
| Linear Regression | Global Cognition, Processing speed, Working memory, SCD, APOE Genotype, Sex, Age at visit | 1.22 | 0.71 |
| SVM | Global Cognition, Processing speed, Working memory, SCD, APOE Genotype, Sex, Age at visit | 0.99 | 0.82 |
| Amyloid Plaques |  |  |  |
| Bagging | Global Cognition, Processing speed, Episodic memory, SCD, APOE Genotype, Sex, Age at visit | 1.16 | 0.69 |
| Gradient Boosting | Global Cognition, Processing speed, Episodic memory, SCD, APOE Genotype, Sex, Age at visit | 1.23 | 0.68 |
| Linear Regression | Global Cognition, Processing speed, Working memory, Episodic memory, SCD, APOE Genotype, Sex | 1.36 | 0.53 |
| SVM | Processing speed, Working memory, Episodic memory, SCD, APOE Genotype, Sex, Age at visit | 1.19 | 0.64 |
| **Amyloid Angiopathy** |  |  |  |
| Bagging | Global Cognition, Processing speed, Working memory, SCD, APOE Genotype, Sex, Age at visit | 0.68 | 0.67 |
| Gradient Boosting | Global Cognition, Processing speed, Working memory, Episodic memory, SCD, APOE Genotype, Sex | 0.62 | 0.74 |
| Linear Regression | Processing speed, Working memory, Episodic memory, SCD, APOE Genotype, Sex, Age at visit | 0.82 | 0.46 |
| SVM | Global Cognition, Working memory, Episodic memory, SCD, APOE Genotype, Sex, Age at visit | 0.78 | 0.54 |
| Abbreviations: WMH = White matter hyperintensity, SCD = Subjective cognitive decline |  |  |  |

In the prediction of diffuse plaques, the Bagging model incorporated Global Cognition, Processing speed, Working memory, Episodic memory, SCD, APOE genotype, and Sex, achieving the lowest RMSE of 0.77 and a correlation of 0.62 (Fig. 1G). The SVM model included a similar set of predictors but focused more on Working memory, recording an RMSE of 0.78 and a correlation of 0.55. The Gradient Boosting model, incorporating Global cognition, Processing speed, Working memory, Episodic memory, SCD, Sex, and Age at visit, demonstrated a higher RMSE of 0.85 and a lower correlation of 0.59. The Linear Regression model, incorporating Global cognition, Processing speed, Working memory, Episodic memory, SCD, APOE genotype, and Age at visit, showed the highest RMSE of 0.89 and the lowest correlation of 0.44.

For neurofibrillary tangles, the Bagging model, including Processing speed, Working memory, Episodic memory, SCD, APOE genotype, Sex, and Age at visit, achieved a correlation of 0.82 and an RMSE of 0.97 (Fig 1H). The SVM model, incorporating the same predictors, had the same correlation of 0.82 but a slightly better RMSE of 0.99. The Gradient Boosting model, including Global cognition, Processing speed, Working memory, Episodic memory, SCD, APOE genotype, Sex, and Age at visit, demonstrated a lower correlation of 0.74 and the highest RMSE of 1.14. The Linear Regression model, incorporating similar predictors, recorded the lowest correlation of 0.71 and an RMSE of 1.22.

In the prediction of amyloid plaques, the Gradient Boosting model, combined Global Cognition, Processing speed, Episodic memory, SCD, APOE genotype, Sex, and Age at visit, exhibited a correlation of 0.68 and an RMSE of 1.23. The Bagging model, including similar predictors, achieved a correlation of 0.69 and an RMSE of 1.16 (Fig 1I). The SVM model, focusing more on Working memory, showed a correlation of 0.64 and an RMSE of 1.19. The Linear Regression model, encompassed Global cognition, Processing speed, Working memory, Episodic memory, SCD, APOE genotype, Sex, and Age at visit, recorded the lowest correlation of 0.53 and the highest RMSE of 1.36.

In predicting amyloid angiopathy, the Gradient Boosting model, combined Global cognition, Processing speed, Working memory, Episodic memory, SCD, APOE genotype, and Sex, achieved the best correlation of 0.74 and the lowest RMSE of 0.62 (Fig. 1J). The Bagging model incorporated Global cognition, Processing speed, Working memory, SCD, APOE genotype, Sex, and Age at visit, with a correlation of 0.67 and an RMSE of 0.68. The SVM model achieved a correlation of 0.54 and an RMSE of 0.78. The Linear Regression model, including Processing speed, Working memory, Episodic memory, SCD, APOE genotype, Sex, and Age at visit, showed the lowest correlation of 0.46 and an RMSE of 0.82.

### Longitudinal neuropathology prediction

To examine whether the ML models are capable of predicting future neuropathology and if MRI features enhance the prognostic predictive performance of the models beyond that achieved with clinical features alone, we conducted additional analysis focusing on longitudinal data across different timepoints prior to death (0-4 years, 4-8 years, and 8-14 years) (see Table 4). The results from the Gradient Boosting model, which was the best performing model overall at predicting neuropathology at last visit are presented for the longitudinal data. Please see the supplementary materials for the results of the other models.

**Table 4:**
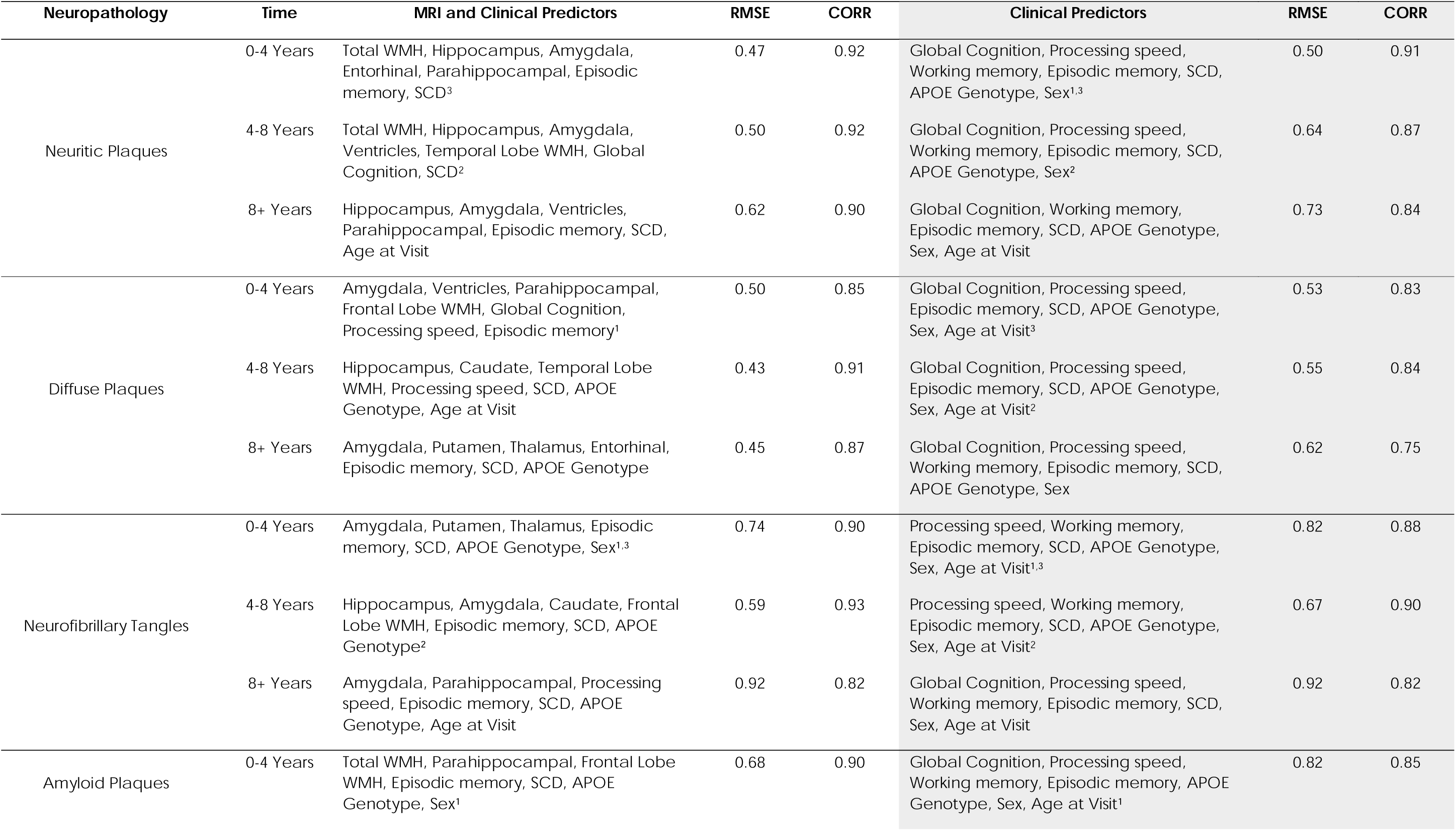
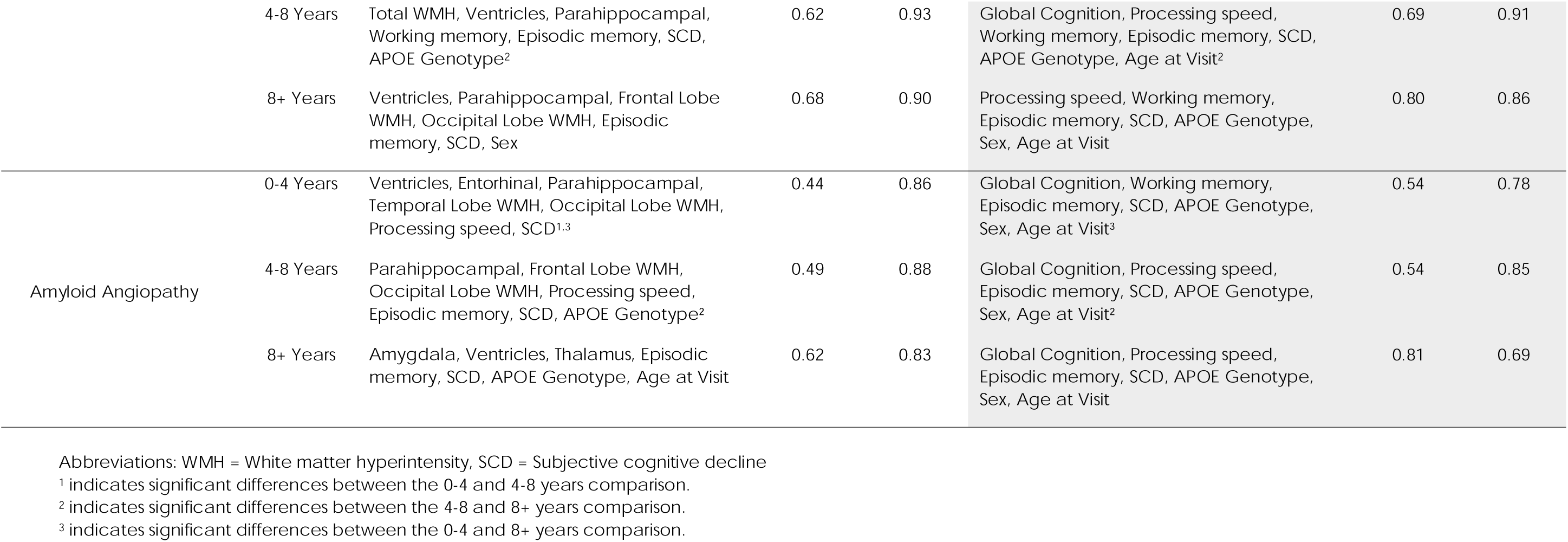
MRI and clinical predictors compared to clinical only predictors of neuropathology outcomes across time when predictors were obtained using the Gradient boosting model.

For the prediction of neuritic plaques between the 0-4 years time prior to death, MRI and clinical predictors such as total WMH, hippocampus, amygdala, entorhinal region, parahippocampal, episodic memory, SCD, and APOE genotype achieved an RMSE of 0.47 and a correlation of 0.92. This model was comparable to the clinical-only model, which achieved an RMSE of 0.50 and a correlation of 0.91. In the 4-8 years interval prior to death, the model utilized total WMH, hippocampus, amygdala, ventricles, temporal lobe WMH, global cognition, and SCD as predictors. The resulting RMSE was 0.50 with a correlation of 0.92, which outperformed the clinical-only model’s RMSE of 0.64 and correlation of 0.87, suggesting enhanced predictive accuracy with the inclusion of MRI data. For the longest interval, 8+ years prior to death, the model incorporated hippocampus, amygdala, ventricles, parahippocampal, episodic memory, and age at visit. The model achieved an RMSE of 0.62 and a correlation of 0.90, surpassing the clinical-only model, which achieved an RMSE of 0.73 and a correlation of 0.84. These results indicate a significant improvement in prediction with the inclusion of MRI features as the time to death increases. For MRI and clinical predictors, small but significant differences were found in the model performances between the 4-8 and 8+ years (*t* = -2.44, *p* = 0.02) and between 0-4 and 8+ years prior to death (*t* = - 2.48, *p* = 0.02). For clinical-only predictors, t-tests demonstrated significant differences at all compared intervals prior to death (*t* values ranging from -8.06 to -2.96, corrected *p*-values < 0.005).

For diffuse plaques, between the 0-4 years prior to death, MRI and clinical predictors such as amygdala, ventricles, parahippocampal, frontal lobe WMH, global cognition, processing speed, and episodic memory achieved an RMSE of 0.50 and a correlation of 0.85. This model was comparable to the clinical-only model, which achieved an RMSE of 0.53 and a correlation of 0.83. In the 4-8 years interval prior to death, the model utilized hippocampus, caudate, temporal lobe WMH, processing speed, SCD, APOE genotype, and age at visit as predictors. The resulting RMSE was 0.43 with a correlation of 0.91, which outperformed the clinical-only model’s RMSE of 0.55 and correlation of 0.84, suggesting enhanced predictive accuracy with the inclusion of MRI data. For the longest interval, 8+ years, the model incorporated amygdala, putamen, thalamus, entorhinal, episodic memory, and SCD. The model achieved an RMSE of 0.45 and a correlation of 0.87, surpassing the clinical-only model, which achieved an RMSE of 0.62 and a correlation of 0.75. For MRI and clinical predictors, t-tests demonstrated significant differences at 0-4 and 4-8 years (*t* = -6.23, *p* < 0.001). For clinical-only predictors, *t*-tests demonstrated significant differences between 4-8 and 8+ years (*t* = -7.10, *p* < 0.001) and between 0-4 and 8+ years (*t* = -10.07, *p* < 0.001).

For neurofibrillary tangles, between the 0-4 years prior to death, MRI and clinical predictors such as the amygdala, putamen, thalamus, episodic memory, SCD, APOE genotype, and sex achieved an RMSE of 0.74 and a correlation of 0.90. This model outperformed the clinical-only model, which achieved an RMSE of 0.82 and a correlation of 0.88. In the 4-8 years interval, the model utilized hippocampus, amygdala, caudate, frontal lobe WMH, episodic memory, SCD, and APOE genotype as predictors. The resulting RMSE was 0.59 with a correlation of 0.93, outperforming the clinical-only model’s RMSE of 0.67 and correlation of 0.90, suggesting enhanced predictive accuracy with the inclusion of MRI data. For the longest interval, 8+ years, the model incorporated amygdala, parahippocampal, processing speed, episodic memory, SCD, APOE genotype, and age at visit. The model achieved an RMSE of 0.92 and a correlation of 0.82, which was identical to the clinical-only model’s RMSE and correlation. This indicates a consistent predictive performance over time. For MRI and clinical predictors and clinical only predictors, *t*-tests demonstrated significant differences at all compared intervals (*t* values ranging from 7.09 to -13.60, corrected *p*-values < 0.001).

For the prediction of amyloid plaques, at the 0-4 years prior to death, MRI and clinical predictors such as total WMH, parahippocampal region, frontal lobe WMH, episodic memory, SCD, APOE genotype, and sex achieved an RMSE of 0.68 and a correlation of 0.90. This model was more accurate than the clinical-only model, which achieved an RMSE of 0.82 and a correlation of 0.85. In the 4-8 years interval, the model utilized total WMH, ventricles, parahippocampal region, working memory, episodic memory, SCD, and APOE genotype as predictors. The resulting RMSE was 0.62 with a correlation of 0.93, outperforming the clinical-only model’s RMSE of 0.69 and correlation of 0.91, indicating improved predictive accuracy with the inclusion of MRI data. For the longest interval, 8+ years, the model incorporated ventricles, parahippocampal region, frontal lobe WMH, occipital lobe WMH, episodic memory, SCD, and sex. The model achieved an RMSE of 0.68 and a correlation of 0.90, which was superior to the clinical-only model’s RMSE of 0.80 and correlation of 0.86. This indicates the consistent predictive advantage of including MRI features. For MRI and clinical predictors, t-tests demonstrated significant differences between 0-4 and 4-8 years (*t* = 10.36, *p* < 0.001) and between 4-8 and 8+ years (*t* = -5.84, *p* < 0.001). For clinical-only predictors, t-tests showed significant differences between 0-4 and 4-8 years (*t* = 4.03, *p* < 0.001) and between 4-8 and 8+ years (*t* = 2.05, *p* = 0.05).

For amyloid angiopathy, between 0-4 years prior to death, MRI and clinical predictors such as the ventricles, entorhinal cortex, parahippocampal region, temporal lobe WMH, occipital lobe WMH, processing speed, and SCD achieved an RMSE of 0.44 and a correlation of 0.86. This model was better than the clinical-only model, which achieved an RMSE of 0.54 and a correlation of 0.78. In the 4-8 years interval prior to death, the model utilized parahippocampal region, frontal lobe WMH, occipital lobe WMH, processing speed, episodic memory, SCD, and APOE genotype as predictors. The resulting RMSE was 0.49 with a correlation of 0.88, outperforming the clinical-only model’s RMSE of 0.54 and correlation of 0.85, indicating enhanced predictive accuracy with the inclusion of MRI data. For the longest interval, 8+ years, the model incorporated amygdala, ventricles, thalamus, episodic memory, SCD, APOE genotype, and age at visit. The model achieved an RMSE of 0.62 and a correlation of 0.83, surpassing the clinical-only model’s RMSE of 0.81 and correlation of 0.69, demonstrating significant improvement in prediction over time. For MRI and clinical predictors, t-tests demonstrated significant differences at all compared intervals (*t* values ranging from 5.72 to -6.85, *p-*values < 0.001). For clinical-only predictors, t-tests showed significant differences between 0-4 and 8+ years (*t*=-5.73, *p*<0.001), and between 4-8 and 8+ years (*t*=-6.94, *p*<0.001). This finding affirms the model’s enhanced predictive capability with the inclusion of MRI features, particularly as the time between MRI follow-up and measurement of pathology (i.e., time to death) increases.

## Discussion

Previous literature has shown the importance of neurofibrillary tangles, amyloid-beta deposits, and alterations in brain morphology, such as GM atrophy and WM abnormalities, in the development of AD [1,3]. Although these studies have recognized the importance of brain changes, the integration of such features into ML models for neuropathology prediction analysis has been less explored. In this study, we investigated the predictive power of ML models in identifying neuropathology outcomes in aging individuals on the spectrum of AD, with a focus on the importance of MRI measures in addition to clinical measures. Our findings reveal that models incorporating MRI predictors, alongside clinical measures demonstrate significant predictive ability for key neuropathological measures such as neurofibrillary tangles, neuritic plaques, diffuse plaques, amyloid plaques, and amyloid angiopathy scores. The longitudinal results indicate a clear trend of increasing number of selected MRI predictors across neuropathology outcomes as the time interval from data collection to death increased, specifically for Neuritic Plaques, Diffuse Plaques, and Amyloid Angiopathy. This is in line with findings from previous studies that investigate the utility of clinical and MRI features in predicting conversion from MCI to dementia in future timepoints [39]. Notably, the models that include MRI features achieved higher performance than models using clinical measures alone at different time intervals prior to death. Gradient boosting, SVM, and bagging models achieved consistently high correlations, gradient boosting however, had superior predictive capabilities, achieving high correlations and low RMSE across most outcomes. When analyzing only clinical predictors, the performance metrics generally indicated lower predictive accuracy for AD neuropathology, reinforcing the added value of incorporating MRI features alongside clinical scores to enhance the predictive capability of ML models. These results are consistent with previous literature highlighting the role of structural and vascular brain changes in cognitive decline and AD progression [2–5]. Results from the present study demonstrate the predictive utility of integrating vascular markers, specifically WMHs, with structural brain changes through ML models.

Our findings also revealed that nonlinear ML models outperformed linear models in predicting AD neuropathology. MRI features do not necessarily influence outcomes in a linear manner; therefore, the nonlinear methods are able to better capture the variability observed in neuropathology. This finding underscores the significance for clinical settings and early prediction research, as it reveals that traditional methods, which assume linear patterns of change, may overlook crucial nuances in neuropathological progression due to the nonlinear influence of predictor variables like GM, WM, WMHs. Nonlinear models, capable of navigating these complexities, can combine information from a set of features to offer more accurate estimation of AD neuropathology and progression than linear models.

The ML models employed in our study exhibited notably high predictive accuracy in estimating neuropathological outcomes as defined by different criteria for neurofibrillary tangles (Braak stage), neuritic and diffuse plaques (CERAD), and amyloid plaques (Thal amyloid phase). In the prediction of neuritic plaques, the SVM model, which used MRI and clinical data, achieved an R² of 0.76, reflecting a strong predictive performance. Comparatively, the SVM model using clinical data only produced an R² of 0.64, highlighting the importance of including MRI data for more accurate predictions even at last visit.

The longitudinal results which incorporate MRI and clinical data from varying time intervals before death (0-4 years, 4-8 years, and 8-14 years), revealed that specific MRI predictors become more useful over time. For instance, regions such as the Amygdala, Frontal Lobe WMH, Hippocampus, Parahippocampal, and Occipital Lobe WMH played increasingly significant roles in long-term predictions. These findings are consistent with the progressive staging of AD. For example, the hippocampus is one of the first regions affected by neurodegeneration[8], and occipital WMHs are the main WMHs associated with AD [4,40]. These findings underscore the importance of utilizing longitudinal MRI data to enhance predictive accuracy.

The comparison of prediction accuracy across time-to-death groups revealed the importance of MRI predictors for future neuropathological outcomes. MRI predictors such as the Amygdala, Hippocampus, and Ventricles remained significant across both last visit and longitudinal analyses, underscoring their critical roles in predicting neuropathological outcomes. For Neurofibrillary Tangles, both datasets identified the Amygdala as an important predictor, although the longitudinal data showed superior predictive accuracy. Additionally, the Frontal and Occipital Lobe WMHs, Ventricles, and Parahippocampal were consistently highlighted as significant predictors in both analyses for outcomes like Amyloid Plaques and Amyloid Angiopathy, respectively.

This high level of predictive performance, achieved using MRI features such as GM, and WMHs, is remarkable, particularly when integrating longitudinal data. The strong performance of these models demonstrates that ML can effectively leverage standard MRI features to provide insights into the underlying neuropathological state, complementing clinical assessments. However, the few studies that have examined neuropathology prediction have been in relatively small samples and report conflicting findings For instance, a study by Toledo et al [41]. utilized cerebrospinal markers, MRI features, and clinical data from 22 ADNI participants and found that non-MRI features were more effective than MRI features in predicting dementia with Lewy bodies in addition to Alzheimer’s pathology. They further highlighted that hippocampal volume was not a reliable feature for predicting pathologies. However, Kautzky et al. [20] applied ML with MRI data in 49 participants to predict neuropathological changes in AD, achieving an accuracy of 77%. Important MRI features highlighted in their study included the entorhinal cortex and anterior cingulate cortex. Building on previous studies, our results underline the potential of integrating advanced machine learning techniques with clinical and MRI data to enhance the predictive accuracy and offer a more comprehensive approach to understanding Alzheimer’s disease progression.

Despite the promising findings of our study, there are limitations that warrant attention. Our study relied on data from the closest MRI timepoint available (under four years from time of death) to maximize sample size. The time difference between the MRI and neuropathology assessments might have impacted the performance of the models. Future studies with closely matched MRI and neuropathology information and longitudinal settings could improve the accuracy of the prediction and assess the change in the prediction accuracy as a function of time between MRI and neuropathology information. The clinical measures and biomarkers used in this study, though carefully selected, do not encompass all possible factors influencing AD neuropathology. Emerging biomarkers and novel imaging techniques continue to be identified, suggesting that integrating these new data sources could potentially enhance the predictive power of future models. Future studies should aim to address these challenges by incorporating longitudinal data and including a broader range of biomarkers to improve prediction accuracy and clinical utility. Lastly, future studies should validate the results in other datasets to ensure generalizability of the selected features. The ability to maintain high performance levels, across multiple dataset’s demographic and clinical diversity, would further emphasize the model’s robust generalizability. This consistency in feature importance across datasets reinforces the reliability and relevance of these features in predicting AD neuropathology, aligning with, and expanding on previous ML research [17,18].

This study combined vascular, structural, and clinical features with machine learning techniques to predict post-mortem neuropathology. We successfully predicted post-mortem neuropathology (∼90% accuracy) in these individuals at up to 14 years before death. Models that included MRI features consistently had the highest accuracy for all AD neuropathology outcomes compared to models with clinical features only. That is, clinical features are successful at predicting AD, but the highest accuracy when predicting AD-related pathology includes MRI features. Not only will these findings improve AD research and patient care in clinical settings, but they could be implemented in clinical trials to select older adults who are at increased risk for AD and who would most benefit from clinical intervention. Our study demonstrates the feasibility and efficacy of using ML models, equipped with a carefully selected set of MRI features, to predict AD neuropathology across diverse cognitive status.

## Data Availability

Data used in preparation of this article were also obtained from the Alzheimers Disease Neuroimaging Initiative (ADNI) database (adni.loni.usc.edu).

## Abbreviations

AD: Alzheimer’s disease
GM: gray matter
WM: white matter
WMH: white matter hyperintensity
MRI: Magnetic resonance imaging
NFT: neurofibrillary tangle burden
ML: machine learning
SVM: support vector machine
ADNI: Alzheimer’s Disease Neuroimaging Initiative
CDR-SB: Clinical Dementia Rating-Sum of Boxes
SCD: subjective cognitive decline
RMSE: root mean square error
FDR: false discovery rate

## Declarations

### Ethics approval and consent to participate

Not applicable.

## Acknowledgments

Data collection and sharing for this project was funded by the Alzheimer’s Disease Neuroimaging Initiative (ADNI) (National Institutes of Health Grant U01 AG024904) and DOD ADNI (Department of Defense award number W81XWH-12-2-0012). ADNI is funded by the National Institute on Aging, the National Institute of Biomedical Imaging and Bioengineering, and through generous contributions from the following: AbbVie, Alzheimer’s Association; Alzheimer’s Drug Discovery Foundation; Araclon Biotech; BioClinica, Inc.; Biogen; Bristol-Myers Squibb Company; CereSpir, Inc.; Cogstate; Eisai Inc.; Elan Pharmaceuticals, Inc.; Eli Lilly and Company; EuroImmun; F. Hoffmann-La Roche Ltd and its affiliated company Genentech, Inc.; Fujirebio; GE Healthcare; IXICO Ltd.; Janssen Alzheimer Immunotherapy Research & Development, LLC.; Johnson & Johnson Pharmaceutical Research & Development LLC.; Lumosity; Lundbeck; Merck & Co., Inc.; Meso Scale Diagnostics, LLC.; NeuroRx Research; Neurotrack Technologies; Novartis Pharmaceuticals Corporation; Pfizer Inc.; Piramal Imaging; Servier; Takeda Pharmaceutical Company; and Transition Therapeutics. The Canadian Institutes of Health Research is providing funds to support ADNI clinical sites in Canada. Private sector contributions are facilitated by the Foundation for the National Institutes of Health (www.fnih.org). The grantee organization is the Northern California Institute for Research and Education, and the study is coordinated by the Alzheimer’s Therapeutic Research Institute at the University of Southern California. ADNI data are disseminated by the Laboratory for Neuro Imaging at the University of Southern California.

## Competing interests

The authors declare no competing interests.

## Funding

This research was supported by a grant from the Canadian Institutes of Health Research (CIHR). Dr. Kamal is supported by a scholarship from Fonds de Recherche du Québec - Santé (FRQS). Dr. Dadar reports receiving research funding from the Quebec Bio-Imaging Network and Fonds de Recherche du Québec - Santé (FRQS), Natural Sciences and Engineering Research Council of Canada (NSERC), Healthy Brains for Healthy Lives (HBHL), Alzheimer Society Research Program (ASRP), CIHR, and Douglas Research Centre (DRC). Dr. Morrison is supported by CIHR.

## Consent Statement

Written informed consent was obtained from participants or their study partner

## Availability of data and materials

Data used in preparation of this article were also obtained from the Alzheimer’s Disease Neuroimaging Initiative (ADNI) database (adni.loni.usc.edu).

## Disclosures

The authors report no disclosures relevant to the manuscript.

## Contributions

F.K, M.O, M.D, and C.M, were involved with the conceptualization and design of the work. F.K. and M.D. completed analysis and C.M, M.D, M.O, and F.K were involved with data interpretation. F.K. organized figures. F.K. wrote the manuscript. and C.M, M.O, M.D, and F.K revised and approved the submitted version.

## Consent for publication

Not applicable.

## Notes

### Competing Interest Statement

The authors have declared no competing interest.

## References

[1] Jack CR, Bennett DA, Blennow K, Carrillo MC, Dunn B, Haeberlein SB, et al. NIA-AA Research Framework: Toward a biological definition of Alzheimer’s disease. Alzheimer’s Dement 2018;14:535–62. 10.1016/j.jalz.2018.02.018.

[2] Bilello M, Doshi J, Nabavizadeh SA, Toledo JB, Erus G, Xie SX, et al. Correlating cognitive decline with white matter lesion and brain atrophy magnetic resonance imaging measurements in Alzheimer’s disease. J Alzheimer’s Dis 2015;48:987–94. 10.3233/JAD-150400.

[3] Dadar M, Manera AL, Ducharme S, Collins DL. White matter hyperintensities are associated with grey matter atrophy and cognitive decline in Alzheimer’s disease and frontotemporal dementia. Neurobiol Aging 2022;111:54–63. 10.1016/j.neurobiolaging.2021.11.007.

[4] Kamal F, Morrison C, Maranzano J, Zeighami Y, Dadar M. Topographical differences in white matter hyperintensity burden and cognition in aging, MCI, and AD. GeroScience 2023;45:1–16. 10.1007/s11357-022-00665-6.

[5] Morrison C, Dadar M, Villeneuve S, Collins DL. White matter lesions may be an early marker for age-related cognitive decline. NeuroImage Clin 2022;35:103096. 10.1016/j.nicl.2022.103096.

[6] Igarashi KM. Entorhinal cortex dysfunction in Alzheimer’s disease. Trends Neurosci 2023;46:124–36. 10.1016/j.tins.2022.11.006.

[7] Fjell AM, McEvoy L, Holland D, Dale AM, Walhovd KB, Initiative. & ADN. What is normal in normal aging? Effects of Aging, Amyloid and Alzheimer’s Disease on the Cerebral Cortex and the Hippocampus. Prog Neurobiol 2014;117:20–40. 10.1016/j.pneurobio.2014.02.004.What.

[8] Risacher SL, Saykin AJ, West JD, Shen L, Firpi HA, Mcdonald BC, et al. Baseline MRI predictors of conversion from MCI to probable AD in the ADNI cohort. Curr Alzheimer Res 2009;6:347–61.

[9] Chen Y, Wang Y, Song Z, Fan Y, Gao T, Tang X. Abnormal white matter changes in Alzheimer’s disease based on diffusion tensor imaging: A systematic review. Ageing Res Rev 2023;87:101911. 10.1016/j.arr.2023.101911.

[10] Van Den Berg E, Geerlings MI, Biessels GJ, Nederkoorn PJ, Kloppenborg RP. White Matter Hyperintensities and Cognition in Mild Cognitive Impairment and Alzheimer’s Disease: A Domain-Specific Meta-Analysis. J Alzheimer’s Dis 2018;63:515–27. 10.3233/JAD-170573.

[11] Gouw AA, Seewann A, Vrenken H, Van Der Flier WM, Rozemuller JM, Barkhof F, et al. Heterogeneity of white matter hyperintensities in Alzheimer’s disease: Post-mortem quantitative MRI and neuropathology. Brain 2008;131:3286–98. 10.1093/brain/awn265.

[12] Piguet O, Double KL, Kril JJ, Harasty J, Macdonald V, McRitchie DA, et al. White matter loss in healthy ageing: A postmortem analysis. Neurobiol Aging 2009;30:1288–95. 10.1016/j.neurobiolaging.2007.10.015.

[13] Bangen KJ, Nation DA, Delano-Wood L, Weissberger GH, Hansen LA, Galasko DR, et al. Aggregate effects of vascular risk factors on cerebrovascular changes in autopsy-confirmed Alzheimer’s disease. Alzheimer’s Dement 2015;11:394–403.e1. 10.1016/j.jalz.2013.12.025.

[14] Di Patre, P. L., Read, S. L., Cummings, J. L., Tomiyasu, U., Vartavarian, L. M., Secor, D. L., & Vinters H V. Progression of clinical deterioration and pathological changes in patients with Alzheimer disease evaluated at biopsy and autopsy. Arch Neurol 1999;56:1254–61.

[15] Iordanishvili E, Schall M, Loução R, Zimmermann, M., Kotetishvili K, Shah NJ, Oros-Peusquens AM. Quantitative MRI of cerebral white matter hyperintensities: a new approach towards understanding the underlying pathology. Neuroimage 2019;202:116077.

[16] Nishiyama Y, Kanayama H, Mori H, Tada K, Yamamoto Y, Katsube T, et al. Whole brain analysis of postmortem density changes of grey and white matter on computed tomography by statistical parametric mapping. Eur Radiol 2017;27:2317–25.

[17] Kaur B, Himali JJ, Seshadri S, Beiser AS, Au R, McKee AC, et al. Association between neuropathology and brain volume in the framingham heart study. Alzheimer Dis Assoc Disord 2014;28:219–25. 10.1097/WAD.0000000000000032.

[18] Zhang M, Ganz AB, Rohde S, Lorenz L, Rozemuller AJM, van Vliet K, et al. The correlation between neuropathology levels and cognitive performance in centenarians. Alzheimer’s Dement 2023;19:5036–47. 10.1002/alz.13087.

[19] Boyle PA, Yu L, Wilson RS, Schneider JA, Bennett DA. Relation of neuropathology with cognitive decline among older persons without dementia. Front Aging Neurosci 2013;5:1–8. 10.3389/fnagi.2013.00050.

[20] Kautzky A, Seiger R, Hahn A, Fischer P, Krampla W, Kasper S, et al. Prediction of autopsy verified neuropathological change of Alzheimer’s disease using machine learning and MRI. Front Aging Neurosci 2018;10:1–11. 10.3389/fnagi.2018.00406.

[21] Mateos-Pérez JM, Dadar M, Lacalle-Aurioles M, Iturria-Medina Y, Zeighami Y, Evans AC. Structural neuroimaging as clinical predictor: A review of machine learning applications. NeuroImage Clin 2018;20:506–22. 10.1016/j.nicl.2018.08.019.

[22] Barnes LL, Shah RC, Aggarwal NT, Bennett DA, Schneider JA. The Minority Aging Research Study: Ongoing efforts to obtain brain donation in African Americans without dementia. Curr Alzheimer Res 2012;9:734–45. 10.2174/156720512801322627.

[23] Coupe P, Yger P, Prima S, Hellier P, Kervrann C, Barillot C. An optimized blockwise nonlocal means denoising filter for 3-D magnetic resonance images. IEEE Trans Med Imaging 2008;27:425–41. 10.1109/TMI.2007.906087.

[24] Sled JG, Zijdenbos AP, Evans AC. A nonparametric method for automatic correction of intensity nonuniformity in mri data. IEEE Trans Med Imaging 1998;17:87–97. 10.1109/42.668698.

[25] Dadar M, Fonov VS, Collins DL. A comparison of publicly available linear MRI stereotaxic registration techniques. Neuroimage 2018;174:191–200. 10.1016/j.neuroimage.2018.03.025.

[26] Fonov V, Evans AC, Botteron K, Almli CR, McKinstry RC, Collins DL. Unbiased average age-appropriate atlases for pediatric studies. Neuroimage 2011;54:313–27. 10.1016/j.neuroimage.2010.07.033.

[27] Dadar M, Maranzano J, Misquitta K, Anor CJ, Fonov VS, Tartaglia MC, et al. Performance comparison of 10 different classification techniques in segmenting white matter hyperintensities in aging. Neuroimage 2017;157:233–49. 10.1016/j.neuroimage.2017.06.009.

[28] Anor CJ, Dadar M, Collins DL, Tartaglia MC. The Longitudinal Assessment of Neuropsychiatric Symptoms in Mild Cognitive Impairment and Alzheimer’s Disease and Their Association With White Matter Hyperintensities in the National Alzheimer’s Coordinating Center’s Uniform Data Set. Biol Psychiatry Cogn Neurosci Neuroimaging 2021;6:70–8. 10.1016/j.bpsc.2020.03.006.

[29] Dadar M, Camicioli R, Duchesne S, Collins DL. The temporal relationships between white matter hyperintensities, neurodegeneration, amyloid beta, and cognition. Alzheimer’s Dement Diagnosis, Assess Dis Monit 2020;12:1–11. 10.1002/dad2.12091.

[30] Dadar M, Maranzano J, Ducharme S, Collins DL. White matter in different regions evolves differently during progression to dementia. Neurobiol Aging 2019;76:71–9. 10.1016/j.neurobiolaging.2018.12.004.

[31] Dadar M, Maranzano J, Ducharme S, Carmichael OT, Decarli C, Collins DL. Validation of T1w-based segmentations of white matter hyperintensity volumes in large-scale datasets of aging. Hum Brain Mapp 2018;39:1093–107. 10.1002/hbm.23894.

[32] Fischl B. Freesurfer. Neuroimage 2012;62:774–81.

[33] Bhattarai P, Taha A, Soni B, Thakuri DS, Ritter E, Chand GB. Predicting cognitive dysfunction and regional hubs using Braak staging amyloid-beta biomarkers and machine learning. Brain Informatics 2023;10:33.

[34] Feurer, M., Klein, A., Eggensperger K, Springenberg JT, Blum M, Hutter F. Auto-sklearn: efficient and robust automated machine learning. Autom Mach Learn 2019:113–34.

[35] Kumari R, Singh J, Gosain A. International Conference on Information and Communication Technology for Intelligent Systems. Singapore Springer Nat Singapore 2023:145–53.

[36] Varoquaux G, Raamana PR, Engemann DA, Hoyos-Idrobo A, Schwartz Y, Thirion B. Assessing and tuning brain decoders: cross-validation, caveats, and guidelines. Neuroimage 2017;145:166–79.

[37] Tekin S, Mega MS, Masterman DM, Chow T, Garakian J, Vinters H V., et al. Orbitofrontal and anterior cingulate cortex neurofibrillary tangle burden is associated with agitation in Alzheimer disease. Ann Neurol 2001;49:355–61. 10.1002/ana.72.

[38] Duchesne S, Caroli A, Geroldi C, Collins DL, Frisoni GB. Relating one-year cognitive change in mild cognitive impairment to baseline MRI features. Neuroimage 2009;47:1363–70. 10.1016/j.neuroimage.2009.04.023.

[39] Zandifar A, Fonov VS, Ducharme S, Belleville S, Collins DL. MRI and cognitive scores complement each other to accurately predict Alzheimer’s dementia 2 to 7 years before clinical onset. NeuroImage Clin 2020;25:102121. 10.1016/j.nicl.2019.102121.

[40] Kaskikallio A, Karrasch M, Koikkalainen J, Lötjönen J, Rinne JO, Tuokkola T, et al. White Matter Hyperintensities and Cognitive Impairment in Healthy and Pathological Aging: A Quantified Brain MRI Study. Dement Geriatr Cogn Disord 2020;48:297–307. 10.1159/000506124.

[41] Toledo JB, Cairns NJ, Da X, Chen K, Carter D, Fleisher A, et al. Clinical and multimodal biomarker correlates of ADNI neuropathological findings. Acta Neuropathol Commun 2014;2:1–13. 10.1186/20515960165.

