## Supplementary Materials for "Exploring the power of MRI and clinical measures in predicting Alzheimer’s disease neuropathology"

**ADNI dataset**

The data used for this article were from the ADNI database (adni.loni.usc.edu), which was established in 2003 as a public-private venture with Michael W. Weiner, MD as the Principal Investigator. ADNI's primary aim is to determine if serial magnetic resonance imaging (MRI), positron emission tomography (PET), other biological markers, and clinical and neuropsychological assessment can be utilized to measure the progression of MCI and AD. The research was granted ethical approval from the review boards of all the involved institutions, and written consent was obtained from participants or their study partner. The participants for this study were taken from all the ADNI cohorts (ADNI-1, ADNI-2, ADNI-GO, and ADNI-3).

**Table 5: MRI and clinical predictors using the Bagging model compared to clinical only predictors of neuropathology outcomes across time when predictors were obtained**

| **Neuropathology** | **Time** | **MRI and Clinical Predictors** | **RMSE** | **CORR** | **Clinical Predictors** | **RMSE** | **CORR** |
| --- | --- | --- | --- | --- | --- | --- | --- |
| Neuritic Plaques | 0-4 Years | Hippocampus, Amygdala, Temporal Lobe WMH, Parietal Lobe WMH, Episodic memory, SCD, APOE Genotype³ | 0.50 | 0.92 | Global Cognition, Processing speed, Working memory, SCD, APOE Genotype, Sex, Age at Visit¹³ | 0.56 | 0.89 |
|  | 4-8 Years | Amygdala, Frontal Lobe WMH, Parietal Lobe WMH, Episodic memory, SCD, APOE Genotype, Sex² | 0.54 | 0.91 | Processing speed, Working memory, Episodic memory, SCD, APOE Genotype, Sex, Age at Visit² | 0.67 | 0.86 |
|  | 8+ Years | Total WMH, Hippocampus, Amygdala, Global Cognition, Episodic memory, SCD, Age at Visit | 0.67 | 0.87 | Global Cognition, Processing speed, Working memory, SCD, APOE Genotype, Sex, Age at Visit | 0.74 | 0.84 |
| Diffuse Plaques | 0-4 Years | Hippocampus, Amygdala, Putamen, Frontal Lobe WMH, Episodic memory, SCD, APOE Genotype | 0.50 | 0.84 | Global Cognition, Processing speed, Episodic memory, SCD, APOE Genotype, Sex, Age at Visit | 0.58 | 0.78 |
|  | 4-8 Years | Total WMH, Ventricles, Putamen, Caudate, Episodic memory, SCD, APOE Genotype | 0.47 | 0.89 | Global Cognition, Working memory, Episodic memory, SCD, APOE Genotype, Sex, Age at Visit | 0.55 | 0.84 |
|  | 8+ Years | Amygdala, Ventricles, Thalamus, Temporal Lobe WMH, Parietal Lobe WMH, Episodic memory, SCD | 0.46 | 0.85 | Global Cognition, Processing speed, Working memory, Episodic memory, APOE Genotype, Sex, Age at Visit | 0.59 | 0.75 |
| Neurofibrillary Tangles | 0-4 Years | Amygdala, Caudate, Thalamus, Processing speed, Episodic memory, SCD, APOE Genotype¹³ | 0.80 | 0.88 | Global Cognition, Processing speed, Working memory, Episodic memory, SCD, APOE Genotype, Age at Visit¹³ | 0.85 | 0.87 |
|  | 4-8 Years | Amygdala, Frontal Lobe WMH, Temporal Lobe WMH, Episodic memory, SCD, APOE Genotype, Sex² | 0.67 | 0.90 | Global Cognition, Processing speed, Working memory, SCD, APOE Genotype, Sex, Age at Visit² | 0.73 | 0.88 |
|  | 8+ Years | Entorhinal, Parahippocampal, Temporal Lobe WMH, Parietal Lobe WMH, Episodic memory, SCD, Age at Visit | 0.99 | 0.79 | Global Cognition, Processing speed, Working memory, Episodic memory, SCD, APOE Genotype, Age at Visit | 0.97 | 0.81 |
| Amyloid Plaques | 0-4 Years | Total WMH, Ventricles, Putamen, Occipital Lobe WMH, Episodic memory, SCD, Age at Visit¹³ | 0.72 | 0.89 | Global Cognition, Processing speed, Working memory, Episodic memory, SCD, APOE Genotype, Age at Visit¹ | 0.86 | 0.83 |
|  | 4-8 Years | Caudate, Parietal Lobe WMH, Processing speed, Working memory, Episodic memory, SCD, APOE Genotype² | 0.61 | 0.93 | Processing speed, Working memory, Episodic memory, SCD, APOE Genotype, Sex, Age at Visit² | 0.71 | 0.91 |
|  | 8+ Years | Entorhinal, Parahippocampal, Frontal Lobe WMH, Temporal Lobe WMH, Episodic memory, SCD, APOE Genotype | 0.68 | 0.89 | Processing speed, Working memory, Episodic memory, SCD, APOE Genotype, Sex, Age at Visit | 0.83 | 0.84 |
| Amyloid Angiopathy | 0-4 Years | Ventricles, Entorhinal, Parahippocampal, Occipital Lobe WMH, Working memory, Episodic memory, SCD³ | 0.46 | 0.84 | Global Cognition, Processing speed, Working memory, Episodic memory, APOE Genotype, Sex, Age at Visit³ | 0.56 | 0.75 |
|  | 4-8 Years | Caudate, Parietal Lobe WMH, Global Cognition, Episodic memory, SCD, APOE Genotype, Age at Visit² | 0.49 | 0.88 | Global Cognition, Processing speed, Episodic memory, SCD, APOE Genotype, Sex, Age at Visit² | 0.54 | 0.85 |
|  | 8+ Years | Thalamus, Parahippocampal, Occipital Lobe WMH, Global Cognition, Episodic memory, SCD, Age at Visit | 0.64 | 0.81 | Global Cognition, Processing speed, Working memory, SCD, APOE Genotype, Sex, Age at Visit | 0.82 | 0.67 |

Abbreviations: WMH = White matter hyperintensity, SCD = Subjective cognitive decline

1 indicates significant differences between the 0-4 and 4-8 years comparison.

2 indicates significant differences between the 4-8 and 8+ years comparison.

3 indicates significant differences between the 0-4 and 8+ years comparison.

**Table 6: MRI and clinical predictors using the SVM model compared to clinical only predictors of neuropathology outcomes across time when predictors were obtained**

| **Neuropathology** | **Time** | **MRI and Clinical Predictors** | **RMSE** | **CORR** | **Clinical Predictors** | **RMSE** | **CORR** |
| --- | --- | --- | --- | --- | --- | --- | --- |
| Neuritic Plaques | 0-4 Years | Amygdala, Ventricles, Thalamus, Parahippocampal, Episodic memory, SCD, APOE Genotype¹³ | 0.56 | 0.90 | Processing speed, Working memory, Episodic memory, SCD, APOE Genotype, Sex, Age at Visit¹³ | 0.59 | 0.88 |
|  | 4-8 Years | Amygdala, Caudate, Temporal Lobe WMH, Occipital Lobe WMH, Episodic memory, SCD, Age at Visit² | 0.66 | 0.87 | Processing speed, Working memory, Episodic memory, SCD, APOE Genotype, Sex, Age at Visit | 0.76 | 0.82 |
|  | 8+ Years | Total WMH, Hippocampus, Parahippocampal, Processing speed, Episodic memory, SCD, Age at Visit | 0.74 | 0.84 | Global Cognition, Processing speed, Working memory, Episodic memory, SCD, APOE Genotype, Age at Visit | 0.74 | 0.84 |
| Diffuse Plaques | 0-4 Years | Hippocampus, Amygdala, Ventricles, Entorhinal, Processing speed, SCD, APOE Genotype¹³ | 0.51 | 0.86 | Global Cognition, Processing speed, Episodic memory, SCD, APOE Genotype, Sex, Age at Visit¹³ | 0.67 | 0.70 |
|  | 4-8 Years | Amygdala, Caudate, Frontal Lobe WMH, Occipital Lobe WMH, Episodic memory, SCD, APOE Genotype² | 0.59 | 0.82 | Global Cognition, Processing speed, Working memory, Episodic memory, SCD, APOE Genotype, Sex² | 0.72 | 0.71 |
|  | 8+ Years | Amygdala, Caudate, Thalamus, Processing speed, Episodic memory, SCD, APOE Genotype | 0.38 | 0.91 | Global Cognition, Processing speed, Working memory, Episodic memory, SCD, APOE Genotype, Age at Visit | 0.51 | 0.81 |
| Neurofibrillary Tangles | 0-4 Years | Hippocampus, Parahippocampal, Working memory, Episodic memory, SCD, APOE Genotype, Age at Visit¹³ | 0.85 | 0.87 | Processing speed, Working memory, Episodic memory, SCD, APOE Genotype, Sex, Age at Visit¹³ | 0.87 | 0.87 |
|  | 4-8 Years | Amygdala, Frontal Lobe WMH, Occipital Lobe WMH, Episodic memory, SCD, APOE Genotype, Age at Visit² | 0.69 | 0.90 | Global Cognition, Processing speed, Working memory, SCD, APOE Genotype, Sex, Age at Visit² | 0.82 | 0.85 |
|  | 8+ Years | Hippocampus, Parahippocampal, Working memory, Episodic memory, SCD, APOE Genotype, Age at Visit | 1.12 | 0.76 | Global Cognition, Processing speed, Working memory, Episodic memory, SCD, APOE Genotype, Age at Visit | 1.17 | 0.72 |
| Amyloid Plaques | 0-4 Years | Amygdala, Caudate, Occipital Lobe WMH, Processing speed, Episodic memory, SCD, APOE Genotype¹³ | 0.94 | 0.81 | Global Cognition, Processing speed, Working memory, Episodic memory, SCD, APOE Genotype, Sex¹³ | 1.00 | 0.77 |
|  | 4-8 Years | Caudate, Thalamus, Frontal Lobe WMH, Occipital Lobe WMH, Episodic memory, SCD, APOE Genotype² | 0.88 | 0.86 | Processing speed, Working memory, Episodic memory, SCD, APOE Genotype, Sex² | 1.09 | 0.77 |
|  | 8+ Years | Amygdala, Ventricles, Occipital Lobe WMH, Global Cognition, Episodic memory, SCD, APOE Genotype | 0.62 | 0.92 | Global Cognition, Processing speed, Episodic memory, SCD, APOE Genotype, Sex, Age at Visit | 0.80 | 0.85 |
| Amyloid Angiopathy | 0-4 Years | Ventricles, Putamen, Parahippocampal, Occipital Lobe WMH, Episodic memory, SCD, APOE Genotype³ | 0.53 | 0.78 | Global Cognition, Working memory, Episodic memory, SCD, APOE Genotype, Sex, Age at Visit¹³ | 0.70 | 0.58 |
|  | 4-8 Years | Ventricles, Thalamus, Parietal Lobe WMH, Episodic memory, SCD, APOE Genotype, Age at Visit² | 0.57 | 0.84 | Global Cognition, Working memory, Episodic memory, SCD, APOE Genotype, Sex² | 0.63 | 0.79 |
|  | 8+ Years | Ventricles, Thalamus, Parietal Lobe WMH, Episodic memory, SCD, APOE Genotype, Age at Visit | 0.68 | 0.79 | Processing speed, Working memory, Episodic memory, SCD, APOE Genotype, Sex, Age at Visit | 0.86 | 0.63 |

Abbreviations: WMH = White matter hyperintensity, SCD = Subjective cognitive decline

1 indicates significant differences between the 0-4 and 4-8 years comparison.

2 indicates significant differences between the 4-8 and 8+ years comparison.

3 indicates significant differences between the 0-4 and 8+ years comparison.

**Table 7: MRI and clinical predictors using the Linear Regression model compared to clinical only predictors of neuropathology outcomes across time when predictors were obtained**

| **Neuropathology** | **Time** | **MRI and Clinical Predictors** | **RMSE** | **CORR** | **Clinical Predictors** | **RMSE** | **CORR** |
| --- | --- | --- | --- | --- | --- | --- | --- |
| Neuritic Plaques | 0-4 Years | Total WMH, Hippocampus, Putamen, Episodic memory, SCD, APOE Genotype, Sex¹ | 0.85 | 0.72 | Global Cognition, Working memory, Episodic memory, SCD, APOE Genotype, Sex, Age at Visit¹ | 0.88 | 0.70 |
|  | 4-8 Years | Amygdala, Putamen, Episodic memory, SCD, APOE Genotype, Sex, Age at Visit² | 0.97 | 0.67 | Global Cognition, Processing speed, Working memory, SCD, APOE Genotype, Sex, Age at Visit² | 0.97 | 0.68 |
|  | 8+ Years | Amygdala, Frontal Lobe WMH, Working memory, Episodic memory, SCD, APOE Genotype, Age at Visit | 0.85 | 0.77 | Processing speed, Working memory, Episodic memory, SCD, APOE Genotype, Sex, Age at Visit | 0.89 | 0.75 |
| Diffuse Plaques | 0-4 Years | Hippocampus, Amygdala, Putamen, Processing speed, Episodic memory, SCD, APOE Genotype¹³ | 0.70 | 0.66 | Global Cognition, Processing speed, Episodic memory, SCD, APOE Genotype, Sex, Age at Visit | 0.80 | 0.52 |
|  | 4-8 Years | Total WMH, Caudate, Thalamus, Temporal Lobe WMH, Episodic memory, SCD, APOE Genotype² | 0.79 | 0.63 | Global Cognition, Processing speed, Working memory, Episodic memory, SCD, APOE Genotype, Sex² | 0.84 | 0.57 |
|  | 8+ Years | Amygdala, Temporal Lobe WMH, Occipital Lobe WMH, Working memory, Episodic memory, SCD, APOE Genotype | 0.61 | 0.72 | Global Cognition, Processing speed, Working memory, Episodic memory, APOE Genotype, Sex, Age at Visit | 0.76 | 0.52 |
| Neurofibrillary Tangles | 0-4 Years | Hippocampus, Putamen, Parahippocampal, Episodic memory, SCD, APOE Genotype, Age at Visit³ | 1.17 | 0.73 | Processing speed, Working memory, Episodic memory, SCD, APOE Genotype, Sex, Age at Visit¹ | 1.23 | 0.70 |
|  | 4-8 Years | Caudate, Temporal Lobe WMH, Global Cognition, Episodic memory, SCD, APOE Genotype, Sex | 1.18 | 0.65 | Global Cognition, Processing speed, Working memory, SCD, APOE Genotype, Sex, Age at Visit² | 1.16 | 0.67 |
|  | 8+ Years | Hippocampus, Putamen, Parahippocampal, Episodic memory, SCD, APOE Genotype, Age at Visit | 1.22 | 0.65 | Global Cognition, Processing speed, Working memory, Episodic memory, SCD, APOE Genotype, Age at Visit | 1.25 | 0.63 |
| Amyloid Plaques | 0-4 Years | Hippocampus, Amygdala, Putamen, Processing speed, Episodic memory, SCD, APOE Genotype¹³ | 1.12 | 0.69 | Global Cognition, Processing speed, Working memory, Episodic memory, APOE Genotype, Sex, Age at Visit | 1.29 | 0.55 |
|  | 4-8 Years | Hippocampus, Amygdala, Caudate, Processing speed, Episodic memory, SCD, APOE Genotype | 1.19 | 0.72 | Global Cognition, Processing speed, Working memory, SCD, APOE Genotype, Sex, Age at Visit | 1.25 | 0.68 |
|  | 8+ Years | Hippocampus, Thalamus, Entorhinal, Parahippocampal, Episodic memory, SCD, APOE Genotype | 1.22 | 0.65 | Global Cognition, Processing speed, Working memory, Episodic memory, APOE Genotype, Sex, Age at Visit | 1.25 | 0.63 |
| Amyloid Angiopathy | 0-4 Years | Hippocampus, Caudate, Parahippocampal, Occipital Lobe WMH, Working memory, Episodic memory, SCD¹³ | 0.66 | 0.63 | Global Cognition, Processing speed, Episodic memory, SCD, APOE Genotype, Sex, Age at Visit³ | 0.79 | 0.39 |
|  | 4-8 Years | Parahippocampal, Occipital Lobe WMH, Working memory, Episodic memory, SCD, APOE Genotype, Age at Visit² | 0.78 | 0.67 | Processing speed, Working memory, Episodic memory, SCD, APOE Genotype, Sex, Age at Visit² | 0.80 | 0.64 |
|  | 8+ Years | Parahippocampal, Occipital Lobe WMH, Working memory, Episodic memory, SCD, APOE Genotype, Age at Visit | 1.01 | 0.42 | Global Cognition, Processing speed, Working memory, Episodic memory, SCD, APOE Genotype, Age at Visit | 1.08 | 0.26 |

Abbreviations: WMH = White matter hyperintensity, SCD = Subjective cognitive decline

1 indicates significant differences between the 0-4 and 4-8 years comparison.

2 indicates significant differences between the 4-8 and 8+ years comparison.

3 indicates significant differences between the 0-4 and 8+ years comparison
